# Coronavirus (COVID-19) infection in children at a specialist centre: outcome and implications of underlying ‘high-risk’ comorbidities in a paediatric population

**DOI:** 10.1101/2020.05.20.20107904

**Authors:** RW Issitt, J Booth, WA Bryant, A Spiridou, AM Taylor, P du Pré, P Ramnarayan, J Hartley, M Cortina-Borja, K Moshal, H Dunn, H Hemingway, NJ Sebire

## Abstract

**Background:** There is evolving evidence of significant differences in severity and outcomes of coronavirus disease 2019 (COVID-19) in children compared to adults. Underlying medical conditions associated with increased risk of severe disease are based on adult data, but have been applied across all ages resulting in large numbers of families undertaking social ‘shielding’ (vulnerable group). We conducted a retrospective analysis of children with suspected COVID-19 at a Specialist Children’s Hospital to determine outcomes based on COVID-19 testing status and underlying health vulnerabilities.

**Methods:** Routine clinical data were extracted retrospectively from the Institution’s Electronic Health Record system and Digital Research Environment for patients with suspected and confirmed COVID-19 diagnoses. Data were compared between Sars-CoV-2 positive and negative patients (CoVPos / CoVNeg respectively), and in relation to presence of underlying health vulnerabilities based on Public Health England guidance.

**Findings:** Between 1^st^ March and 15^th^ May 2020, 166 children (<18 years of age) presented to a specialist children’s hospital with clinical features of possible COVID-19 infection. 65 patients (39.2%) tested positive for SARS-CoV-2 virus. CoVPos patients were older (median 9 [0.9 - 14] years vs median 1 [0.1 - 5.7.5] years respectively, *p*<0.001). There was a significantly reduced proportion of vulnerable cases (47.7% vs 72.3%, *p*=0.002), but no difference in proportion of vulnerable patients requiring ventilation (61% vs 64.3%, *p* = 0.84) between CoVPos and CoVNeg groups. However, a significantly lower proportion of CoVPos patients required mechanical ventilation support compared to CoVNeg patients (27.7 vs 57.4%, *p*<0.001). Mortality was not significantly different between CoVPos and CoVNeg groups (1.5 vs 4% respectively, *p*=0.67) although there were no direct COVID-19 related deaths in this highly preselected paediatric population.

**Interpretation:** COVID-19 infection may be associated with severe disease in childhood presenting to a specialist hospital, but does not appear significantly different in severity to other causes of similar clinical presentations. In children presenting with pre-existing ‘COVID-19 vulnerable’ medical conditions at a specialist centre, there does not appear to be significantly increased risk of either contracting COVID-19 or severe complications, apart from those undergoing chemotherapy, who are over-represented.

## Introduction

The 2019 novel coronavirus SARS-CoV-2 causes potentially severe respiratory and gastrointestinal symptoms in humans (coronavirus disease; COVID-19).^1^ Infection may be asymptomatic or clinically manifest as mild coryzal symptoms through to pneumonia, severe acute respiratory distress syndrome, multi-organ failure and death.^2,3^ As of the 20^th^ May 2020, there have been almost 5 million confirmed cases of COVID-19, and >300,000 associated deaths.^1^ The major forms of human coronavirus disease (SARS, MERS and COVID-19) can infect children but appear to be associated with fewer symptoms and less severe disease compared with adults, with correspondingly lower case-fatality rates.^2,4-6^

In relation to the current COVID-19 pandemic, children are commonly infected but are more likely to be either asymptomatic or develop only mild non-specific symptoms, although rarely severe respiratory complications may occur. Children also present more frequently than adults with gastrointestinal symptoms, although persistent fever is reported, which is uncommon with other novel coronaviruses.^2,5,6^

In the United Kingdom, Public Health England (PHE) have outlined a shielding strategy designed to protect those within the population deemed to be “extremely vulnerable” to COVID-19 infection.^7,8^ This includes individuals that are immunocompromised, have specific types of cancer, severe respiratory conditions and other rare diseases. For these groups, based on adult data, the recommendation is to remain isolated in their homes for several months, with significant impact on the individual and their families. Given the documented reduced overall severity reported in children with COVID-19, we examined a cohort of paediatric patients presenting to a specialist children’s hospital with suspected COVID-19 to document their clinical behaviour and outcomes with particular regard to presence of underlying medical conditions leading to “vulnerability”.

## Methods

### Study design and participants

Retrospective analysis of routinely collected hospital data fulfiling the ethical guidelines of the Helsinki Declaration and approved by the Institutional Review Board as part of a wider study for the use of routine hospital data within a secure digital research environment (17/LO/0008). All clinical data was collated in a research platform within the hospital’s governance structure and de-identified prior to analysis. Data from patients presenting to a specialist children’s hospital in London, UK between 1^st^ March and 15^th^ May 2020 with a suspected diagnosis of COVID-19 were included.

### Data Collection

Routine clinical information was extracted from the Institution’s Electronic Health Record system using a custom structured query language script. These data included patient demographic information, laboratory results, admission and hospital stay information, intensive care requirements and outcome. Clinical outcomes were as of 15^th^ May 2020. COVID-19 positive cases (CoVPos) were defined as patients presenting with clinical features of possible COVID-19, based on attending clinician interpretation of published features including fever, cough and systemic symptoms, with either direct positive molecular test results documented at the referring or presenting hospital (positive for SARSCoV-2 by real-time PCR test for nucleic acid in respiratory or blood samples), or patients with similar clinical features associated with documented positive familial COVID-19 testing. COVID-19 negative cases (CoVNeg) were defined as those presenting with clinical features of possible COVID-19 but with documented negative COVID-19 testing. Patients were classified regarding vulnerability group using the “COVID-19 – high risk shielded patient list identification methodology” published by NHS Digital, (which was augmented by the Institution’s local policy to include specific paediatric subgroups such as congenital immunodeficiencies).^8^ For seasonal comparison, de-identified hospital admission data, including vulnerability status, was extracted for the calendar year 2019.

As part of the hospital policy, patients during the study period were tested for Sars-Cov-2 based on clinical suspicion of possible COVID-19, including working diagnoses of infection/sepsis, flu-like illness and respiratory tract infection. CoVPos patients were treated according to the relevant specialty guides for patient management during the coronavirus pandemic, either based on National Health Service or specialty specific guidance.

### Statistical Analysis

Continuous variables were summarised by their median and interquartile range. Categorical variables were described by counts and percentages. Proportional testing of groups was undertaken using a 2-sample test for equality of proportions with continuity correction. Analysis of differences between groups was undertaken using 2-tailed *t*-tests or WilcoxonMann-Whitney tests according to the variables’ distribution. Linear mixed effects regression models with a random effect to account for within-patient measurements were fitted to analyse laboratory measurements. COVID-19 diagnosis and vulnerable status (without interaction term) were used as fixed effects. P-values were obtained by likelihood ratio tests of the null model with the random effect against the model with COVID-19 diagnosis and COVID-19 diagnosis plus vulnerability status. Statistical tests were conducted assuming a 0.05 significance level. All analyses were performed in the R language and environment for statistical computing, version 3.5.0 (R Foundation for Statistical Computing, Vienna, Austria).

## Results

### Demographic and clinical characteristics

A total of 166 patients presented to the specialist children’s hospital with suspected diagnosis of COVID-19 during the period between 1^st^ March and 15th May 2020 (Figure 1). Sixty five patients were confirmed COVID-19 positive (CoVPos; 39.2%; median age 9 years (range 1 week – 17.2 years)). 101 patients were COVID-19 negative (CoVNeg; 60.8%; median age 1 years (range 1 day – 16.7 years), who were significantly younger than the CoVPos group (*p*<0.001). Both groups were similar in ethnic backgrounds and gender (Table 1, Figure 2) although those of Asian background were overrepresented in the CoVPos group (18.5 vs 5.9%; *p* = 0.02). The most common diagnosis codes for the CoVNeg group were sepsis, septic shock, fever and pneumonia; CoVPos patients were diagnosed as COVID-19 disease.

**Figure 1.**
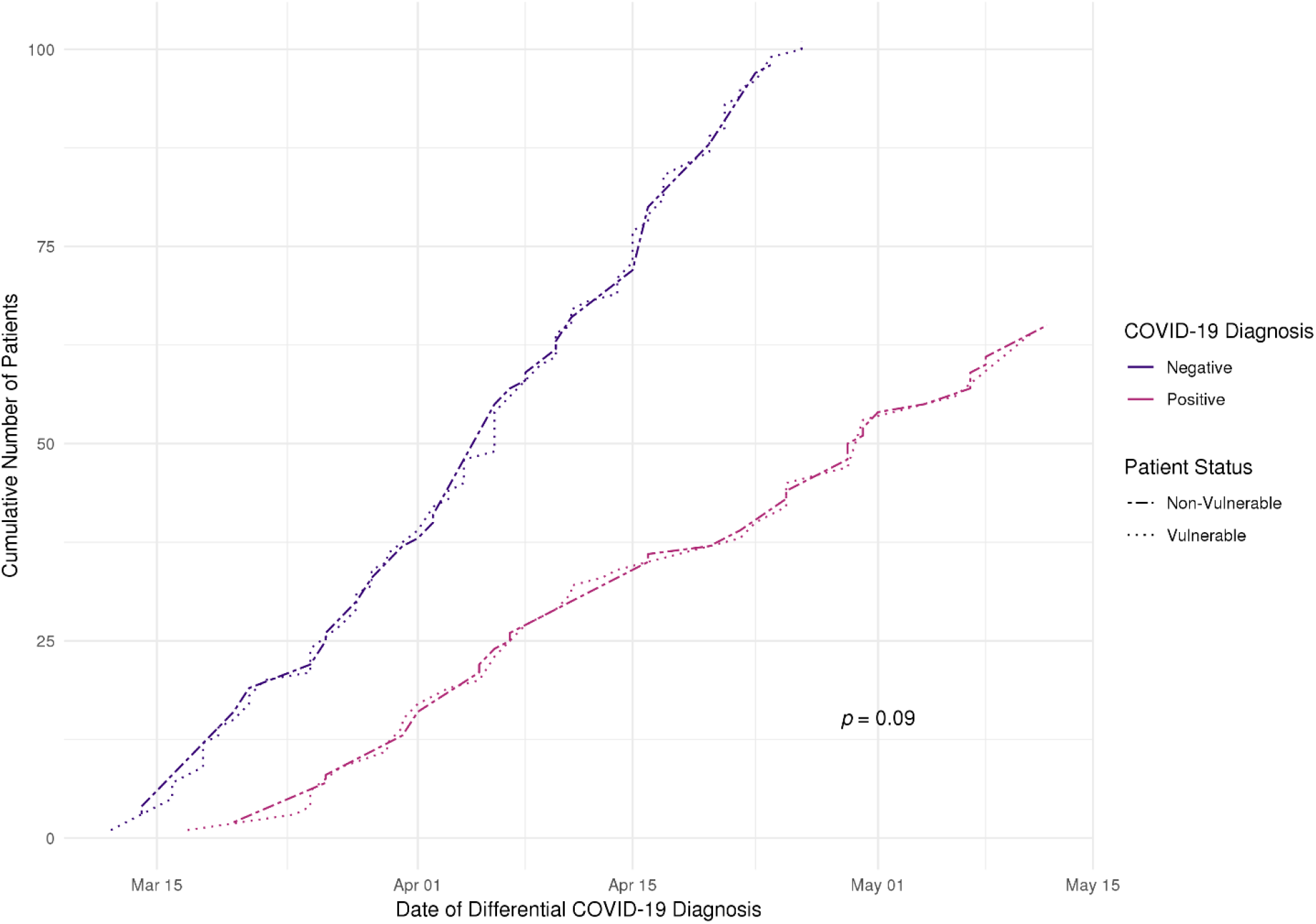
Presentations to a specialist children’s hospital faceted by COVID-19 diagnosis, and whether classed as vulnerable due to underlying conditions. Vulnerable status was not related to rate of presentation or CoVPos diagnosis (slope analysis *p*=0.09).

**Table 1.** Patient demographic data, proportion in vulnerable groups and mortality of 168 CoVPos and CoVNeg patients presenting to a specialist children’s hospital with symptoms suggestive of COVID-19 disease.

|  | COVID-19<br>Negative | COVID-19<br>Positive | *p* |
| --- | --- | --- | --- |
| **Number of Patients** | 101 (60.8) | 65 (38.7) |  |
| **Male** | 58 (55.4) | 38 (58.5) | 0.82 |
| **Female** | 45 (44.6) | 27 (41.5) | 0.82 |
| **Age (Years)** | 1 [0.1 - 5.75] | 9 [0.9 - 14] | **<0.001** |
| **Ethnicity** |  |  |  |
| **Asian** | 6 (5.9) | 12 (18.5) | **0.02** |
| **Black African** | 6 (5.9) | 6 (9.2) | 0.59 |
| **Other** | 4 (4) | 3 (4.6) | 1 |
| **Refused** | 66 (65.3) | 31 (47.7) | **0.035** |
| **White** | 19 (18.9) | 13 (20) | 0.94 |
| **Vulnerable\*** | 73 (72.3) | 31 (47.7) | **0.002** |
| **Mortality** | 4 (4) | 1 (1.5)** | 0.67 |
| **Mortality in Vulnerable** | 4 (4) | 1 (1.5) | 0.67 |
| **Mortality in Non-Vulnerable** | 0 | 0 | 1 |

\*Based on UK guidance for vulnerable groups<sup>8</sup>
\*\*The death in this group occurred in a child with proven SARS-CoV-2 infection but was related to pre-existing underlying conditions and associated coinfections.

**Figure 2.**
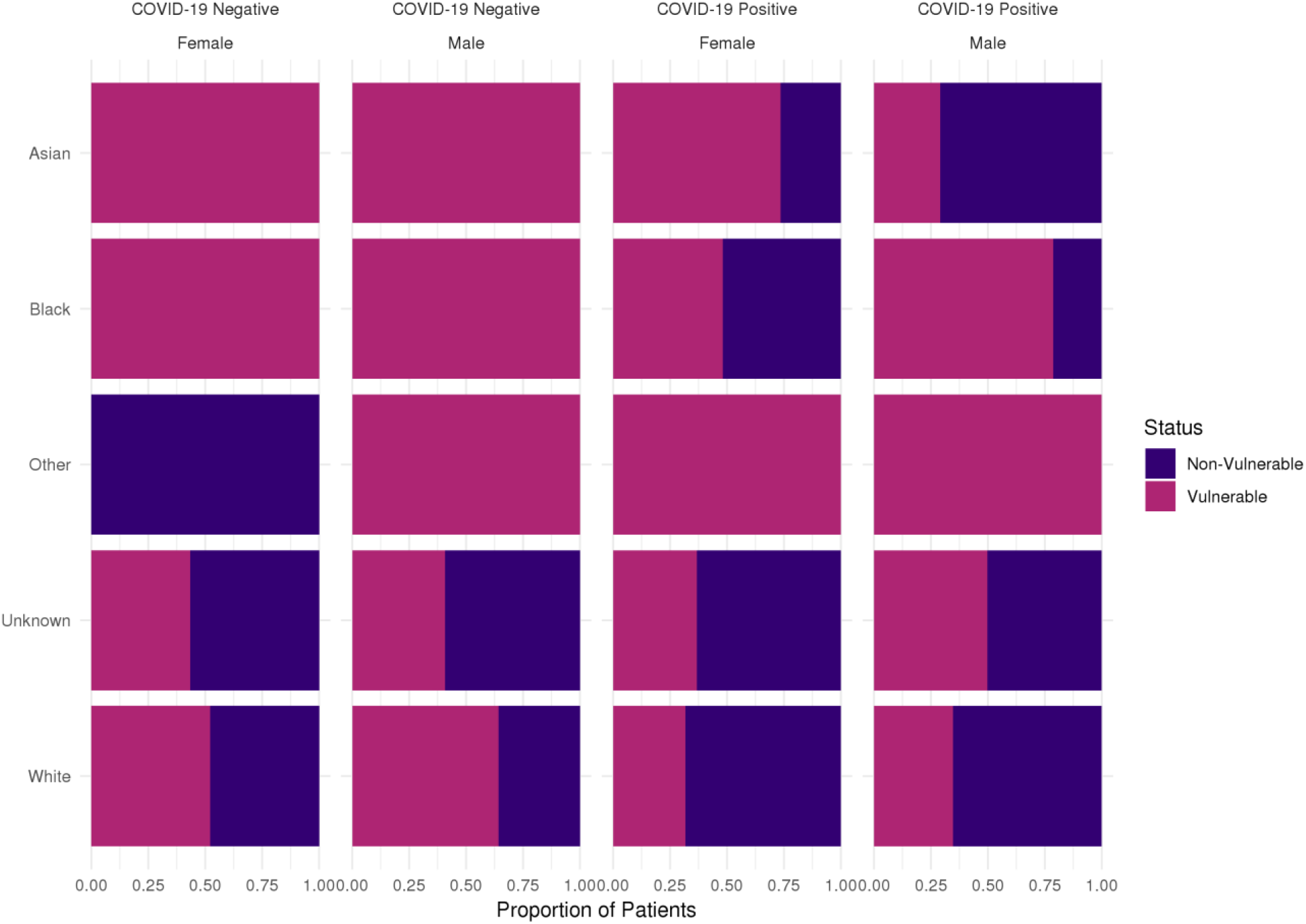
Ethnic background for 166 paediatric symptomatic patients presenting with suspected COVID-19 disease to a specialist children’s hospital faceted by COVID-19 diagnosis. (Unknown includes those patients whose parents preferred not to say and where no ethnicity was recorded).

There was a significantly lower proportion of patients classed as vulnerable in the CoVPos (47.7%) versus CoVNeg (72.3%) groups (*p*=0.002; Figure 3). Mortality was similar between CoVPos and CoVNeg groups (1 vs 4; 1.5 vs 4% respectively; *p* = 0.67), all of whom were classed as vulnerable in both groups.

**Figure 3.**
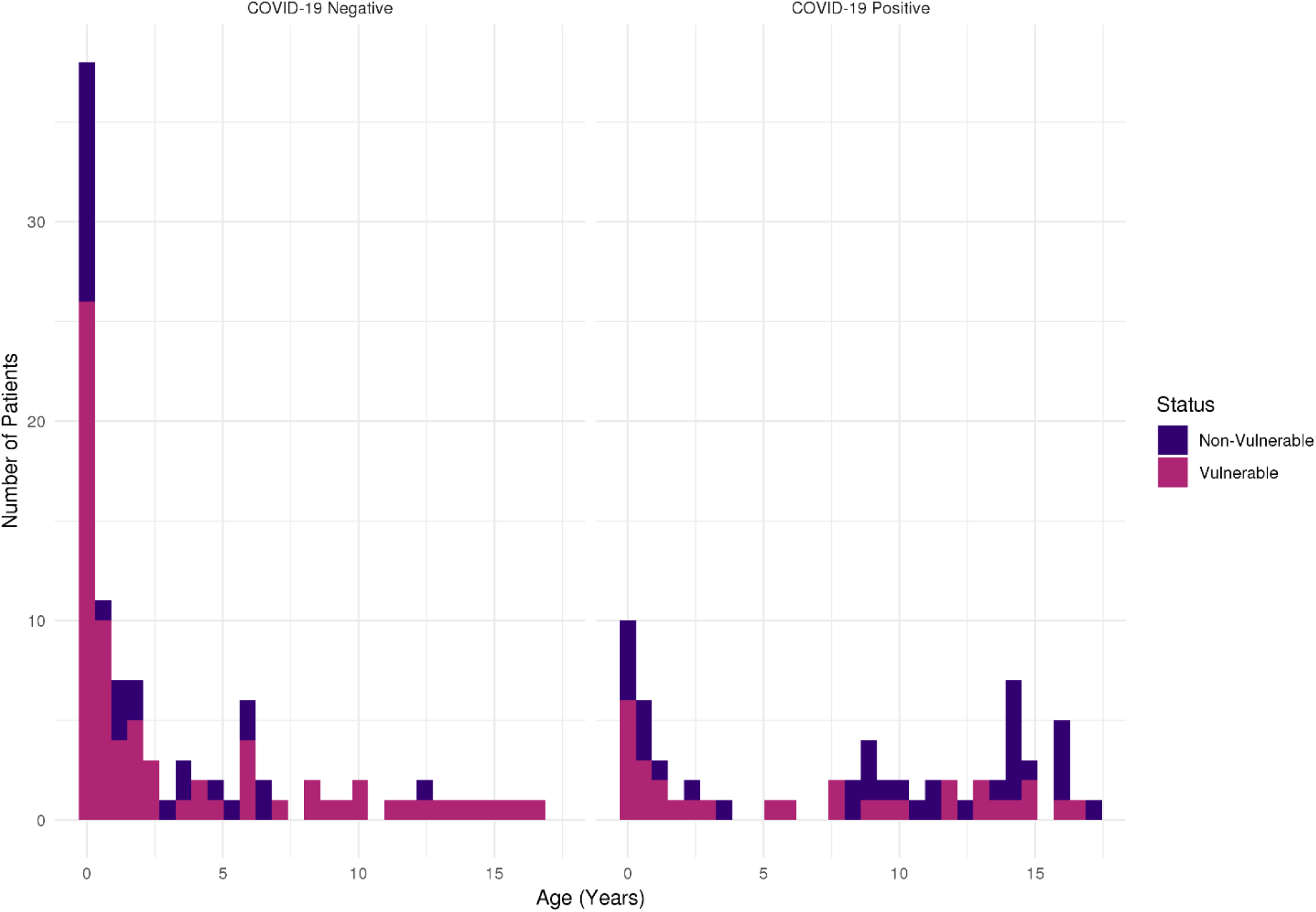
Age and vulnerability distribution for 166 paediatric patients presenting with suspected COVID-19 disease.

The CoVPos patient that died had confirmed SARS-CoV-2 infection but had other underlying severe health conditions and died from aspiration pneumonia with SARS-CoV-2 infection likely representing either an incidental feature or indirect contributor to death. There were no cases of direct COVID-19 related death in this paediatric series in vulnerable or non-vulnerable patients. The four patients in the CoVNeg group died of other causes including sepsis and bacterial meningitis.

Analysis of hospital specialty admissions demonstrated that paediatric intensive care and general paediatrics were the largest recipients of patients overall. The majority of patients (n=60 and 27; CoVNeg and CoVPos respectively) were emergently transferred from outlying hospitals. (Figure 4).

**Figure 4.**
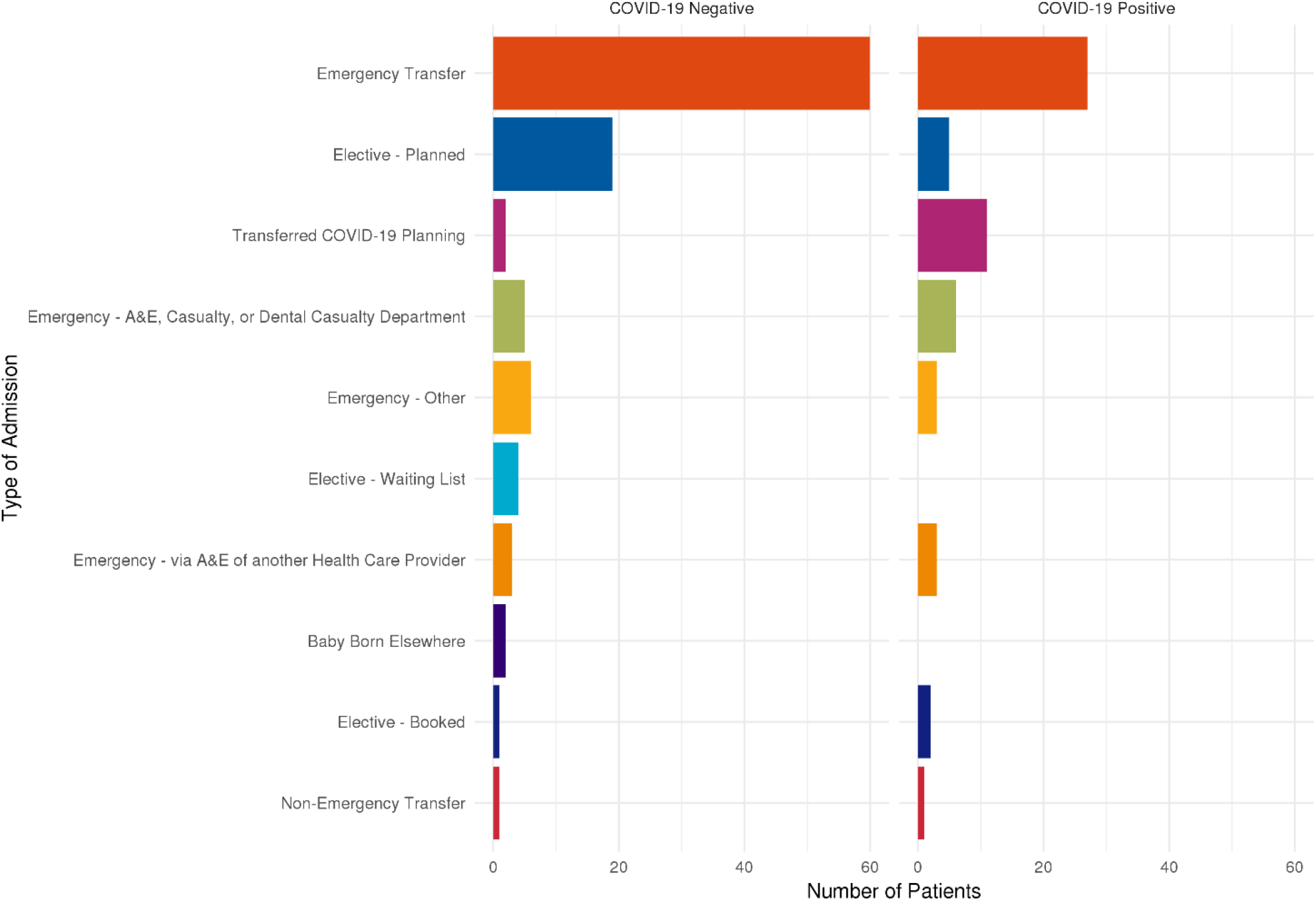
Presentation type for 166 paediatric patients symptomatic with suspected COVID19 disease.

During the study period, with a daily average of 326 inpatients, on average 10 were CoVPos at any time, representing around 3% of the hospital inpatient population: compared to around 25% COVID-19+ inpatient proportion across adult London trusts.^9^

### Laboratory Results

A total of 19,670 laboratory results were generated among all patients. Between-group analysis of laboratory tests undertaken showed similar patterns of testing for both cohorts (Figure 5). Using a linear mixed effects model to examine the impact of COVID-19 diagnosis, there were significant differences in Albumin (CoVNeg 29 [25–34] g/L vs CoVPos 32 [27–36] g/L, p=0.02), C-reactive protein (22 [8–51] ml/L vs 28 [10–74] mg/L, p=0.002), Fibrinogen (2.4 [1.8–3.7] g/L vs 3.65 [2.4–4.8] g/L; *p*<0.001) and Lactate Dehydrogenase (923 [764–2106] U/L vs 848 [654–1136] U/L, p=0.002). The addition of vulnerability status to the mixed effects model showed significance in both Neutrophil and White Blood Cell counts (p<0.001; Table 3).

**Table 2.** Proportion of patients (N (%)) by vulnerable underlying disease group in children presenting with symptoms and tested for COVID-19. There was no overall difference in the proportion of vulnerable patients between groups although patients undergoing cancer chemotherapy are significantly over-represented.

|  | COVID-19<br>Negative | COVID-19<br>Positive | *p* |
| --- | --- | --- | --- |
| **1 (National) Transplant** | 6 (8.2) | 1 (3.2) | 0.62 |
| **1d (National) Transplant Medication** | 4 (5.5) | 1 (3.2) | 1 |
| **2a (National) Cancer undergoing active chemo/radiotherapy** | 3 (4.1) | 7 (22.6) | **0.01** |
| **2b (National) Haematological Cancers** | 2 (2.7) | 2 (6.4) | 0.73 |
| **3 (National) Respiratory** | 2 (2.7) | 1 (3.2) | 1 |
| **4 (National) Rare genetic, metabolic and autoimmune diseases** | 7 (9.6) | 5 (16.2) | 0.54 |
| **C (Local) People with severe respiratory conditions** | 21 (29) | 5 (16.2) | 0.27 |
| **D (Local) People with rare diseases** | 5 (6.8) | 1 (3.2) | 0.79 |
| **E (Local) People on immunosuppression therapies** | 1 (1.4) | 1 (3.2) | 1 |
| **O (Local) Other potential factors** | 22 (30) | 7 (22.6) | 0.58 |

**Table 3.** Laboratory results for CoVNeg and CoVPos paediatric patients. Linear mixed effects models were used to compare random effects of individual patients to additional fixed effects of COVID-19 diagnosis (model 1) and COVID-19 diagnosis and vulnerability status (model 2). Data are presented as grouped median [IQR] values across admission.

|  | COVID-19<br>Negative | COVID-19<br>Positive | *p*<br>Model 1 | *p*<br>Model 2 |
| --- | --- | --- | --- | --- |
| Alanine Transaminase (U/L) | 31 [23 – 54] | 41.5 [29 – 74] | 0.52 | 0.99 |
| Albumin (g/L) | 29 [25 – 34] | 32 [27 – 36] | **0.02** | 0.99 |
| AntiDNAase B (U/mL) | 69.6 [69.6 – 508] | 310 [80.8 – 402] | 0.58 | 0.88 |
| AntiStreptolysin O (IU/mL) | 64.4 [12.7 – 317] | 285 [134 – 384] | 0.31 | 0.82 |
| Aspartate Transaminase (U/L) | 56 [34 – 76] | 70 [43 – 100] | 0.34 | 0.89 |
| C-Reactive Protein (mg/L) | 22 [8 – 51] | 28 [10 – 74] | **0.002** | 0.49 |
| Creatine Kinase (U/L) | 409 [96.5 – 895] | 63.5 [35 – 214] | 0.09 | 0.82 |
| Creatinine (μmol/L) | 24 [15 – 52] | 23 [14 – 46] | 0.31 | 0.38 |
| D-Dimers (μg/L) | 1964 [582 – 4514] | 1876 [1043 – 3618] | 0.13 | 0.92 |
| Ferritin (μg/L) | 397 [284 – 650] | 788 [445 – 1863] | 0.5 | 0.36 |
| Fibrinogen (g/L) | 2.4 [1.8 – 3.7] | 3.65 [2.4 – 4.8] | **<0.001** | 0.71 |
| Interleukin-6 (pg/mL) | 50 [50 – 50] | 50 [50 – 152] | 0.4 | 0.73 |
| Interleukin-10 (pg/mL) | 50 [50 – 50] | 50 [50 – 50] | 0.58 | 0.30 |
| Lactate Dehydrogenase (U/L) | 923 [764 – 2106] | 848 [654 – 1136] | **0.002** | 0.22 |
| Lymphocytes (x10<sup>9</sup>/L) | 1.71 [0.75 – 3.25] | 1.44 [0.64 – 2.49] | 0.25 | 0.36 |
| Neutrophils (x10<sup>9</sup>/L) | 3.34 [1.68 – 6.84] | 3.90 [1.46 – 8.60] | 0.34 | **<0.001** |
| NT-pro-Brain Natriuretic Peptide (pg/mL) | 9004 [6522 – 21736] | 3550 [626 – 6992] | 0.06 | 0.95 |
| Prothrombin time (seconds) | 12.1 [11.4 – 13.5] | 12 [11.3 – 13] | 0.2 | 0.83 |
| Total bilirubin (μmol/L) | 9 [4 – 21] | 6 [3 – 10] | 0.053 | 0.40 |
| Triglycerides (mmol/L) | 1.85 [1.43 – 2.72] | 2.48 [1.65 – 3.56] | 0.37 | 0.31 |
| Troponin I (ng/L) | 147 [50 – 310] | 54 [13 – 157] | 0.11 | 0.42 |
| Urine Total Protein (g/L) | 0.13 [0.11 – 0.51] | 0.12 [0.12 – 0.62] | 0.72 | 0.61 |
| White Blood Cells (x10<sup>9</sup>/L) | 7.47 [3.15 – 11.8] | 8 [3.38 – 13.2] | 0.73 | **<0.001** |

**Figure 5.**
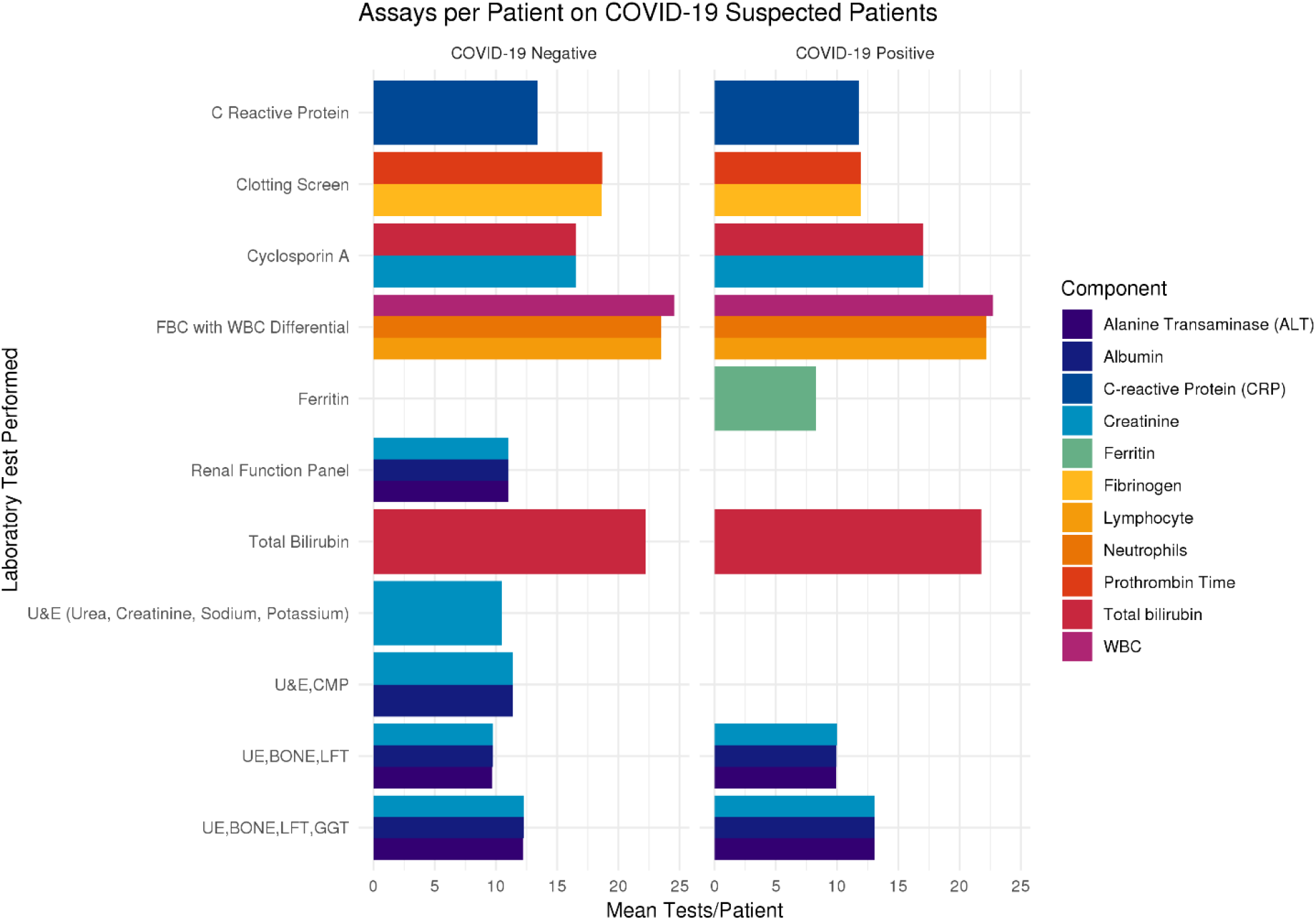
Most common laboratory tests for 166 paediatric patients symptomatic with suspected COVID-19 disease showing similar investigation profile.

### Vulnerable patients

Overall, vulnerable patients were not over-represented in the CoVPos group, comprising 47.7% of the cohort. However, subanalysis of the vulnerable patient cohort revealed significant over-representation of vulnerable patients in specific national coding group 2(a) “Cancer undergoing active chemo/radiotherapy” in the CoVPos cohort (22.6% vs 4.1%, *p* = 0.01; Figure 6). All other vulnerable groups showed no significant differences between groups (*p* = 0.27-1; Table 2).

**Figure 6.**
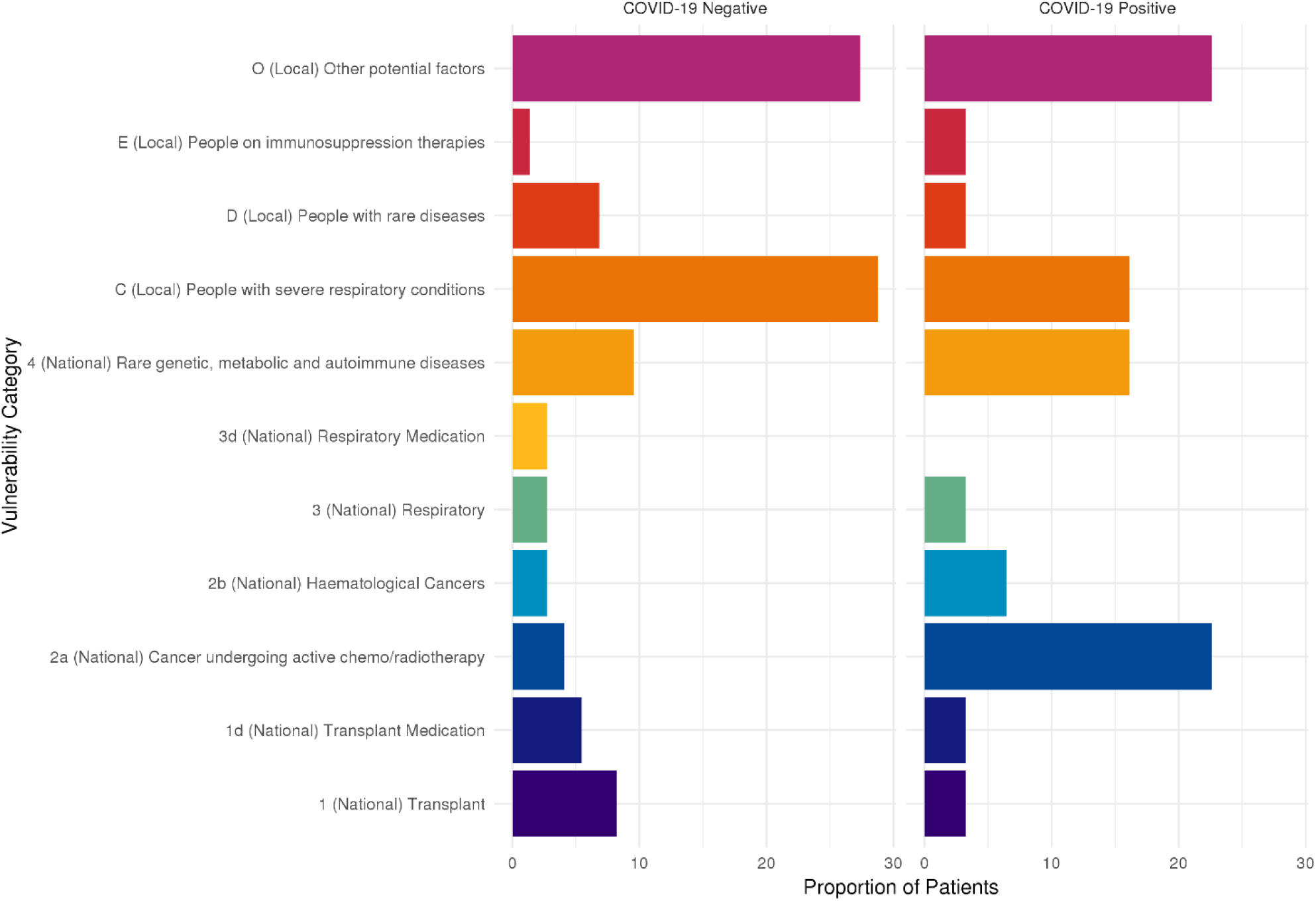
Vulnerability groups for admissions of 166 paediatric patients symptomatic with suspected COVID-19 disease. There was no overall difference in the proportion of vulnerable patients between groups, although patients undergoing cancer chemotherapy (2a) are significantly over-represented in CoVPos group (*p* = 0.003).

The overall proportion of vulnerable patients admitted to the hospital during the COVID-19 emergency study period was compared to historical data from the 2019. The proportion of patients from vulnerable groups observed across March to May in the CoVPos cohort was 47.7% versus 72.3% in the CoVNeg cohort (*p*=0.002). During 2019 the mean proportion of patients classed as vulnerable within the hospital was 79.3% for inpatients and 47.5% for outpatients. Proportional testing showed a significantly lower proportion of vulnerable inpatients (*p* 0.001). The CoVNeg cohort was not significantly different to the background inpatient proportion in terms of vulnerability (72.3% vs 79.3%, *p* = 0.17).

Comparison of patients requiring intensive care and respiratory support (defined as the requirement for mechanical ventilation), demonstrated a significantly lower proportion of CoVPos patients requiring respiratory support compared to CoVNeg patients presenting with similar clinical features (27.7% vs 57.4%, *p*<0.001)/ There was no difference between groups in the proportion of vulnerable patients requiring respiratory support (61% vs 64.3%, respectively, *p*=0.84, Figure 7).

**Figure 7.**
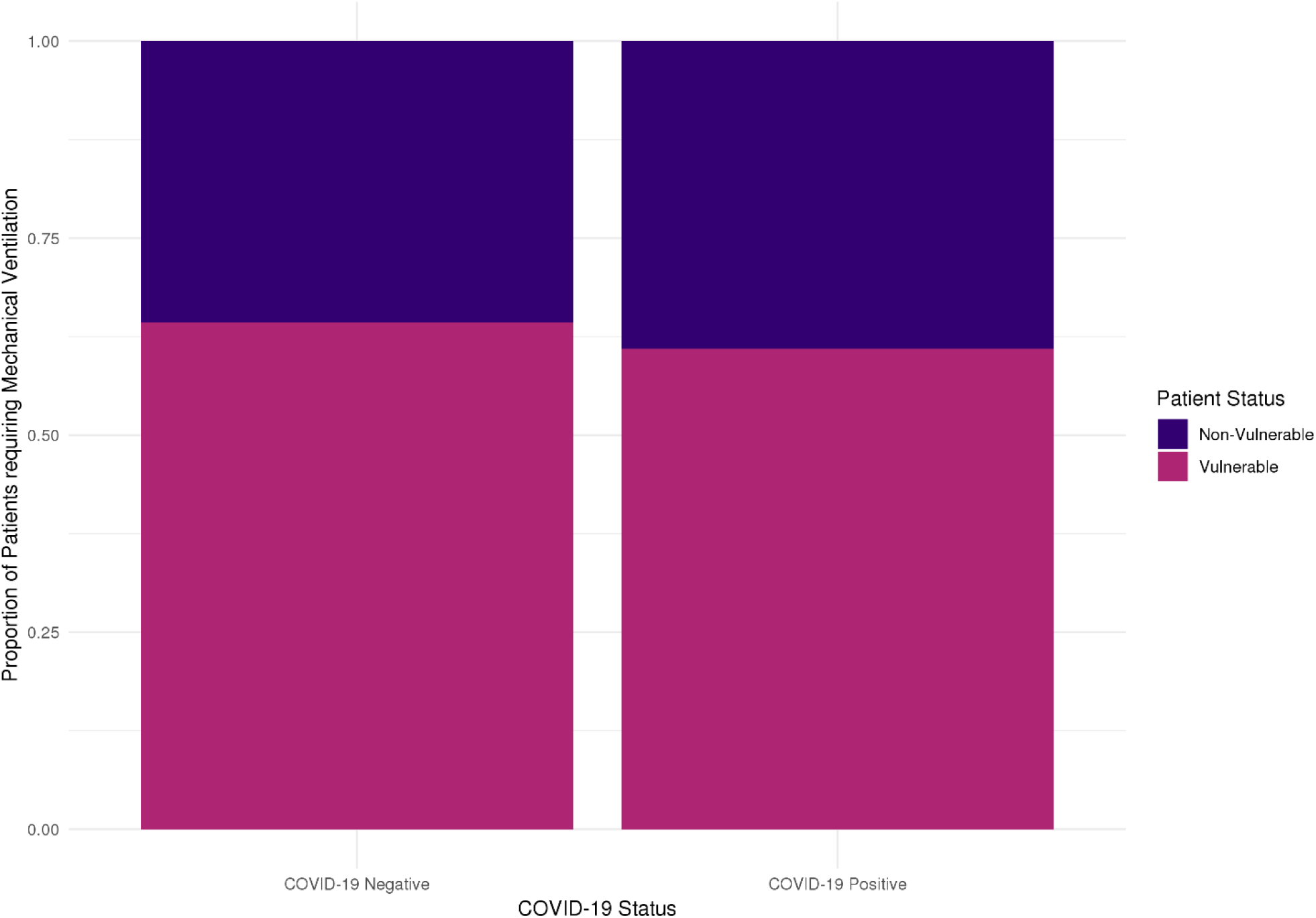
Patient requirement for mechanical ventilator support for 166 paediatric patients symptomatic with suspected COVID-19 disease. The proportion of patients requiring mechanical ventilation was not significantly increased in the CoVPos group (*p* = 0.6).

As of 15^th^ May, there were 10 CoVPos and 23 CoVNeg patients still hospitalised (9 and 13 in intensive care respectively). Length of stay on intensive care was not significantly different between CoVPos and CoVNeg groups (*p*=0.46) and was also similar when vulnerability was accounted for (*p*=0.3, Figure 8). Overall hospital stay was significantly shorter in the CoVPos than CoVNeg group (3.94 [2.5–15.7] days vs 9.1 [4.1–18.9] days respectively, *p*=0.01) and was also not significantly different for vulnerable patients between groups (*p*=0.94, Figure 9, Table 4).

**Figure 8.**
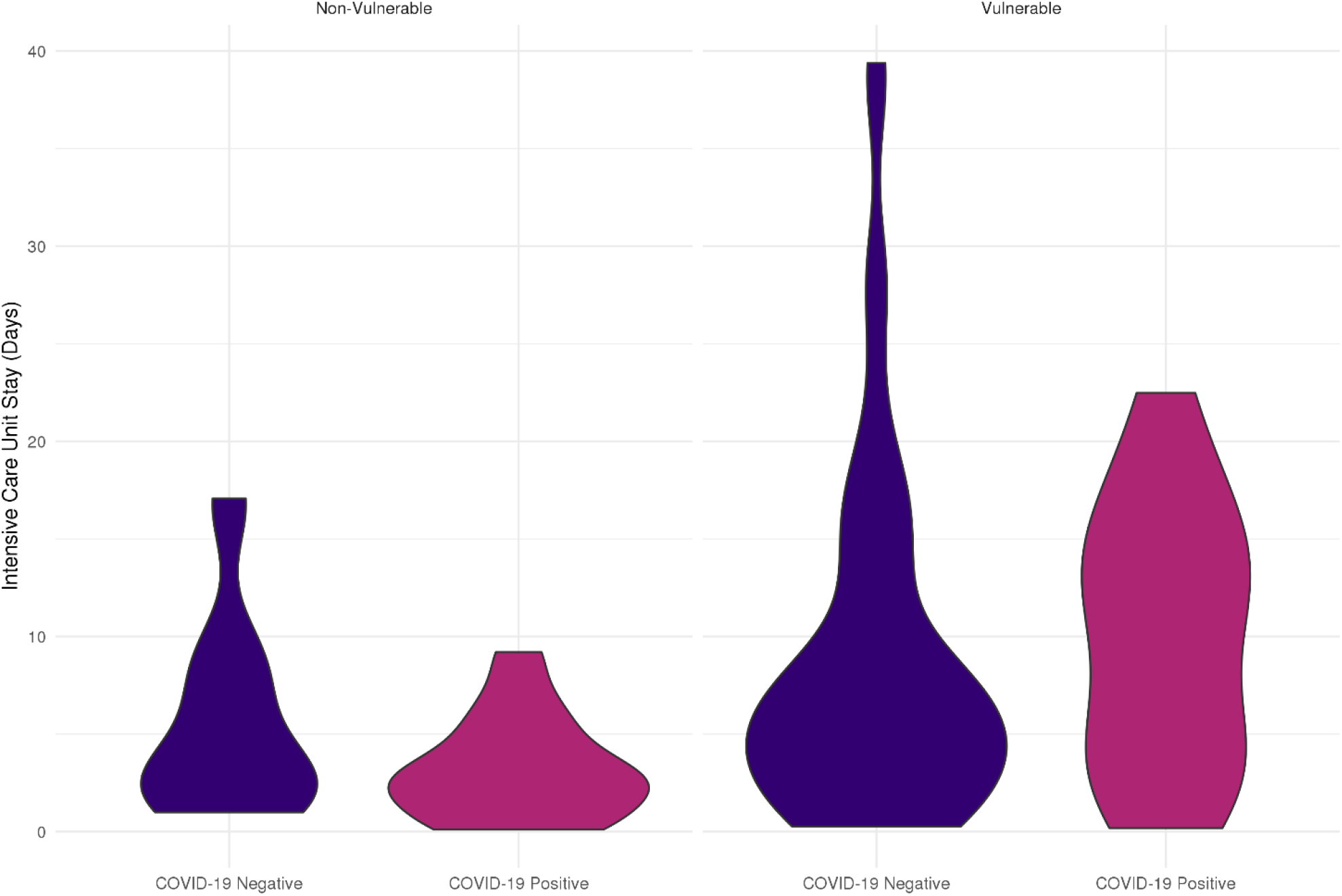
Overview of Intensive Care Stay for paediatric patients symptomatic with suspected COVID-19 disease. There was no significant increase in length of intensive care stay for CoVPos patients.

**Figure 9.**
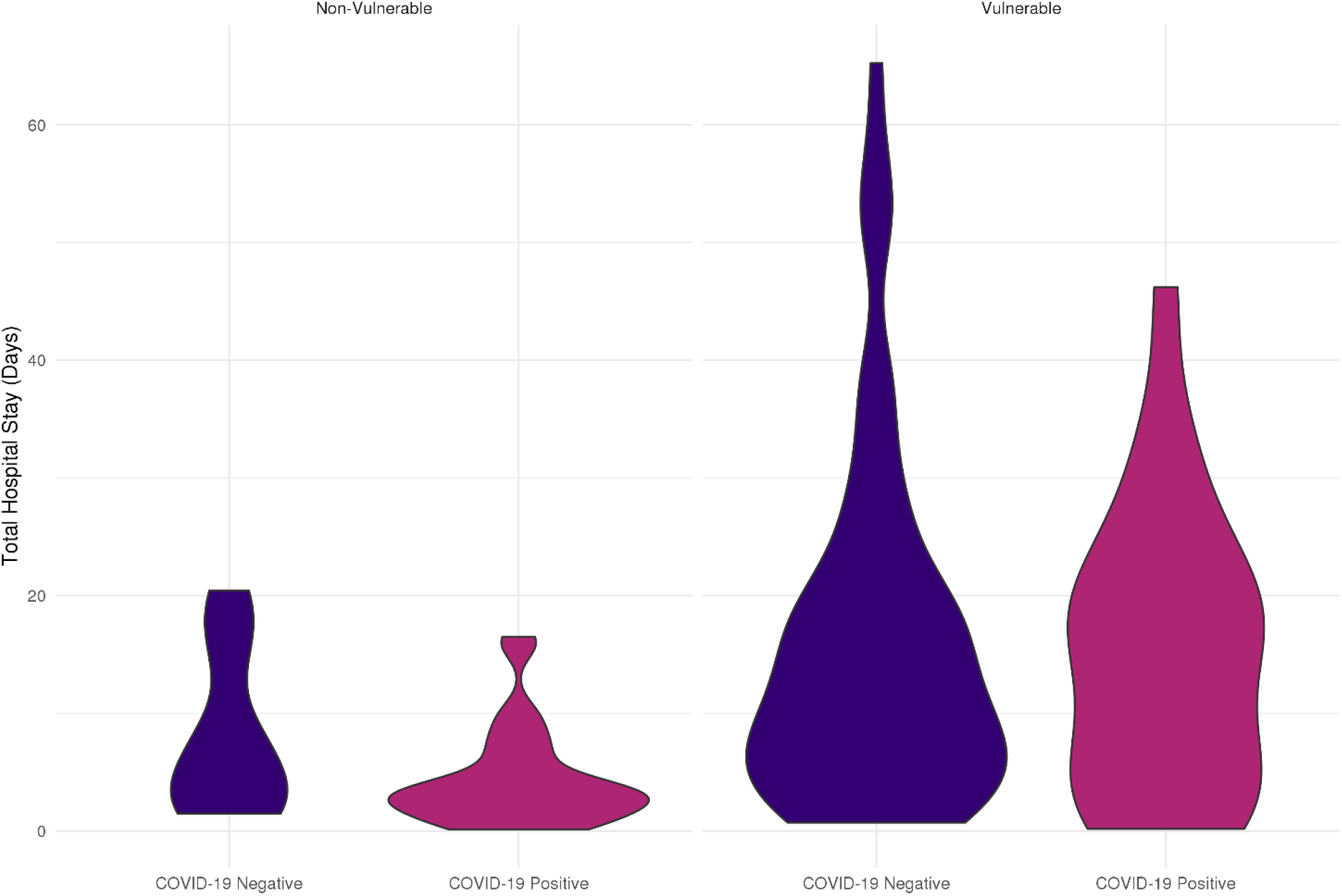
Overview of total hospital stay for paediatric patients symptomatic with suspected COVID-19 disease. There was no significant increase in overall length of stay for CoVPos patients.

**Table 4.**
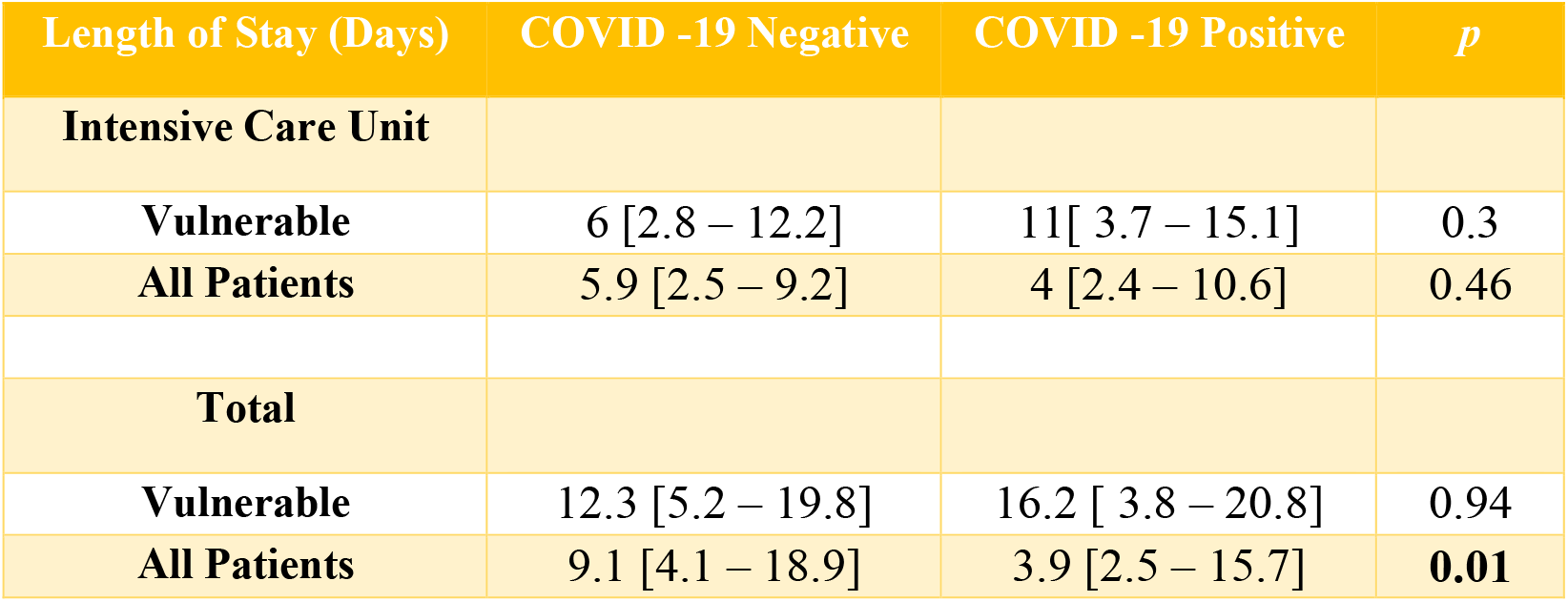
Length of hospital and ICU stay data of CoVPos and CoVNeg paediatric patients by vulnerability status.

| Length of Stay (Days) | COVID -19 Negative | COVID -19 Positive | *p* |
| --- | --- | --- | --- |
| **Intensive Care Unit** |  |  |  |
| **Vulnerable** | 6 [2.8 – 12.2] | 11[ 3.7 – 15.1] | 0.3 |
| **All Patients** | 5.9 [2.5 – 9.2] | 4 [2.4 – 10.6] | 0.46 |
| **Total** |  |  |  |
| **Vulnerable** | 12.3 [5.2 – 19.8] | 16.2 [ 3.8 – 20.8] | 0.94 |
| **All Patients** | 9.1 [4.1 – 18.9] | 3.9 [2.5 – 15.7] | **0.01** |

## Discussion

The findings of this study have demonstrated the characteristics and outcome of children presenting to a highly specialist children’s hospital with clinical features suggestive of COVID-19 disease. In this context, approximately forty percent of patients tested positive for SARS-CoV-2 confirming COVID-19 disease, while the remainder had a negative SARS-Cov-2 test. In some patients it should be noted that their presentation to hospital may have been a direct result of severe underlying illnesses, and that the finding of COVID-19 appeared to be incidental. Testing for COVID-19 in these children was due to clinical suspicion of possible COVID-19 with working diagnoses of sepsis or other causes of flu-like illnesses / respiratory tract infections, and because of the high prevalence in the population. In paediatric patients presenting with similar clinical features, CoVPos patients were not significantly more likely to be admitted to the intensive care unit or require mechanical ventilation and there was no difference in overall mortality between the CoVPos and CoVNeg groups. These data suggest that, whilst in children COVID-19 may occasionally present as severe disease, it does not appear to be had an adverse clinical course compared to other causes of similar presentation.

The findings of the current study confirm that children may be infected with SARS-CoV-2 and this may lead to severe disease with requirement for intensive care admission and mechanical ventilation, and/or other intensive support in rare cases. It should be recognised that as a specialist children’s hospital, the cohort of patients included in the present study are highly preselected, both for children with the most severe disease and those with known underlying medical conditions placing them in the ‘vulnerable’ group, and therefore the findings presented here are not applicable to an unselected paediatric cohort. Previous data from general centres suggests that less than 1% of all of admissions due to COVID-19 represent children under 18 years of age and a recent study from multiple centres in the United States reported only few COVID19 positive patients per hospital intensive care unit.^6,10^ Nevertheless, the data from the current series indicate that COVID-19 represents an additional cause of children being admitted to hospital with severe systemic and/or respiratory disease.

As a highly specialist regional and national children’s hospital, the underlying cohort of patients will significantly and disproportionately represent those with complex underlying medical conditions, hence meeting criteria for ‘vulnerability’. Indeed, around two thirds of patients registered with the hospital in 2019 would be considered as potentially vulnerable according to local interpretation of UK government guidance on underlying conditions for COVID-19.^8^ This is not unexpected, since the centre represents a large unit for paediatric transplantation, rare genetic diseases such as congenital immunodeficiency and treatment for paediatric malignancy. However, a striking feature of the data presented here is the finding that the overall proportion of patients considered vulnerable is not significantly increased in those with confirmed COVID-19 disease, compared to either those with similar presentations but testing negative, or indeed the underlying background prevalence of medical vulnerability conditions among these preselected patients. Furthermore, in those with confirmed COVID-19 disease, the proportion of patients with underlying vulnerable conditions requiring intensive care admission for mechanical ventilation and length of time admitted to intensive care were not increased. There were no deaths directly attributable to COVID-19. These data broadly suggest that, in contrast to the data from adults, the majority of underlying medical conditions do not appear to place children at significantly increased risk of either developing COVID-19 disease or experiencing severe symptoms and complications if infected. Recent data suggests that those adults who are immunocompromised, for example subsequent to renal transplantation, demonstrate increased severe complication rates with COVID-19 (28% mortality).^11^ In contrast, a recent study reported no mortality in a multicenter cohort of patients with cystic fibrosis affected by COVID-19,^12^ and therefore the susceptibility among vulnerable groups is likely to be both disease-specific and related to patient age. In the present study for example, for the majority of underlying conditions there was no difference between the groups, although children undergoing chemotherapy for paediatric malignancy were over-represented in the CoVPos group, suggesting these specific patients are at increased risk of Sars-Cov-2 infection. Further work is therefore required to elucidate the specific conditions associated with increased risk and their associations with age, since this will have significant implications for societal shielding and future strategies to manage lockdowns.

In addition to the typical features of COVID-19 disease described in adults, whilst most children who are infected appear to have mild disease,^2,13^ there has been a recent suggestion that a small minority of children presenting with such clinical features represent an unusual associated systemic inflammatory condition, (paediatric inflammatory multisystem syndrome temporally related to Sars-Cov-2 infection), regardless of their SARS-CoV-2 test result.^14^ At present the criteria for the definitive diagnosis of such a potential syndrome remain undetermined, and it is therefore uncertain whether any of the patients described in this series, including those in the CoVNeg group, could represent such a disorder. In order to determine whether the suggested presentation represents a true increase in frequency associated with the COVID-19 pandemic, further data is required across national paediatric intensive care units, and such studies are ongoing.^14^

This study represents a large, single centre cohort from a specialist children’s hospital, including all patients clinically suspected of presenting with COVID-19 disease. However, the study has limitations due to its retrospective nature, lack of ‘normal’ control group being available for the patients studied, and separation by COVID-19 status without matching. This is evident in the significantly different age demographic of the two cohorts. Furthermore, as diagnostic codes classed as vulnerable will change as more data becomes available, patients classed as non-vulnerable in this study, may yet be considered vulnerable in the future and vice-versa. The main limitation is that the group represents a highly preselected population in a specialist children’s hospital of patients presenting with severe disease and therefore is not representative of the unselected paediatric population as a whole. It should also be noted that since vulnerable children are shielding at home, the pattern of presentation reported here may not be representative of a non-shielded situation. Nevertheless, these data represent a relatively large series of children presenting with similar clinical features and as such provides important data regarding the disease severity of COVID-19 in children compared to other similar infectious conditions and the potential impact of underlying medical condition vulnerability in this population. In particular, the current policy of extended shielding for vulnerable patients of all ages, represents a significant burden for patients and their families in terms of social interactions, schooling and education and childhood mental health. These data suggest that the criteria for vulnerability derived from initial adult evidence may not be applicable to children to the same extent. Further large-scale data are urgently required to understand the implications of underlying disease for COVID-19 outcomes by age in order to develop rational and evidenced based policies for management of children with these conditions in relation to ongoing COVID-19 emergency policy and lockdown management.

In summary, the findings of this study have reported on a cohort of patients presenting to a specialist children’s hospital with clinical features suggestive of COVID-19 infection, in whom approximately forty percent of cases tested positive for the disease. Compared to patients with similar presentation who tested negative, children with COVID-19 do not appear to be at significantly increased risk of severe complications and in particular, in the paediatric population, those with underlying vulnerable conditions do not appear to be greatly at risk of severe disease.

## Data Availability

Deidentified aggregated data is available on request from the corresponding author

## Declaration of interests

The authors declare no conflicts of interests.

## Funding

RI is funded by a British Heart Foundation Research Fellowship Grant. HH is funded by NIHR UCLH BRC and HDRUK, NJS is funded by GOSHCC and HDRUK.

## Role of the funding source

The study sponsor / funders had no role or influence in study design, in the collection, analysis, and interpretation of data, in the writing of the report or in the decision to submit the paper for publication

## Role of the authors

The corresponding author (RI) and NJS,JB, confirm that they had full access to all the data in the study and had final responsibility for the decision to submit for publication. NJS, RI and AT conceived the study. JB, RI, MC, and WB performed the analyses. All authors contributed to the critical appraisal and writing of the manuscript and approved the final submission.

## Research in context

### Evidence before this study

Published evidence to date identified through searching PubMed/Web of Science with the key words ((children OR childhood OR pediatric OR paediatric) AND COVID-19) report that children with COVID-19 appear to be less severely affected than adults. Vulnerable individuals have been identified based on adult data regarding underlying health conditions (NHS Digital).

### Added value of this study

These data report on a highly preselected group of high risk paediatric patients attending a specialist children’s hospital with features suggestive of COVID-19, and demonstrate that around one third of such cases represent confirmed acute infection with SARS-CoV-2. However, the proportion of patients with underlying health conditions rendering them vulnerable was not significantly increased in this population compared to the background population for the hospital, and there was no significant difference in outcome between children who tested positive or negative for SARS-CoV-2. The findings suggest that typical factors representing vulnerable patients identified from adult evidence may not have similar importance for children in relation to COVID-19, with implications for public health shielding approaches. Appropriate identification of high risk groups in childhood is required.

## Notes

### Competing Interest Statement

The authors have declared no competing interest.

